# A phenome-wide association study of tandem repeat variation in 168,554 individuals from the UK Biobank

**DOI:** 10.1101/2024.01.22.24301630

**Authors:** Celine A. Manigbas, Bharati Jadhav, Paras Garg, Mariya Shadrina, William Lee, Alejandro Martin-Trujillo, Andrew J. Sharp

## Abstract

Most genetic association studies focus on binary variants. To identify the effects of multi-allelic variation of tandem repeats (TRs) on human traits, we performed direct TR genotyping and phenome-wide association studies in 168,554 individuals from the UK Biobank, identifying 47 TRs showing causal associations with 73 traits. We replicated 23 of 31 (74%) of these causal associations in the All of Us cohort. While this set included several known repeat expansion disorders, novel associations we found were attributable to common polymorphic variation in TR length rather than rare expansions and include *e.g.* a coding polyhistidine motif in *HRCT1* influencing risk of hypertension and a poly(CGC) in the 5’UTR of *GNB2* influencing heart rate. Causal TRs were strongly enriched for associations with local gene expression and DNA methylation. Our study highlights the contribution of multi-allelic TRs to the “missing heritability” of the human genome.

## Introduction

Over the past two decades, thousands of genome-wide association studies (GWAS) have been performed to identify genetic variants that impact diverse human traits^1^. However, the almost universal reliance of GWAS on genotypes of bi-allelic single nucleotide variants (SNVs) creates certain limitations. Modern GWAS typically perform relatively sparse genotyping of common SNVs and then infer additional genotypes through statistical imputation, a process that relies on patterns of linkage disequilibrium (LD) within the genome. As a result, a fundamental limitation of SNV-based GWAS is that they have reduced ability to assay genetic variants that have low levels of LD with flanking SNVs. Accordingly, complex genomic loci that may include multi-allelic variants that undergo recurrent or high mutation rates are typically poorly assayed by GWAS, a fact which is believed to contribute to the so-called “missing heritability problem”^2^.

One class of variant that has been proposed as a candidate to explain the missing heritability of the genome are tandem repeats (TRs)^3,4^. TRs represent a diverse class of sequence element characterized by multiple tandem copies of a DNA motif repeated in a head-to-tail fashion, *e.g.* CAG-CAG-CAG, with ∼1 million of these loci scattered throughout the human genome. TRs are found within coding, genic and intergenic regions and comprise a wide diversity of motif sizes, ranging from single nucleotide repeats, *e.g.* poly(A), at one extreme, up to large macrosatellites at the other that are composed of repeated motif units that can be several kilobases in size^5,6^. TRs represent some of the most variable parts of the human genome, with some showing extremely high levels of length polymorphism and mutation rates that are typically several orders of magnitude higher than that of SNVs^7^. Paradoxically, until the advent of large-scale genome sequencing and specialized bioinformatic tools^8–12^, TRs were relatively poorly studied, often being neglected due to a lack of technologies able to genotype them at scale.

While extreme expansions of short tandem repeats (STRs) have been known for >30 years as a cause of rare neurodegenerative and congenital disorders^13^, recent studies have shown that common length polymorphism of both STRs and TRs with larger motif sizes can exert functional effects on the genome, contributing to inter-individual differences in gene expression, splicing and DNA methylation^14–20^. Studies of large TRs have demonstrated that variation in these can also modify human phenotypes^21,22^ and, for some diseases, TRs represent the strongest known common genetic risk factors^23^. However, to date, only a single published study has attempted to assess the global contribution of STR variation with a small number of human traits^24^. To address this knowledge gap, here we leveraged available genome sequencing data to genotype a set of the most polymorphic human TRs with motif sizes ranging from 2-20bp in >168,000 individuals from the UK Biobank (UKB) and used these to perform a phenome-wide association study (PheWAS), identifying dozens of TRs that causally influence human traits.

## Results

### A phenome-wide association study of TRs in the UK Biobank

We identified a set of 48,913 TRs with (i) motif sizes ranging from 2-20bp, (ii) were either highly polymorphic or showed evidence of rare expansion and (iii) preferentially overlapped either coding regions, UTRs and regulatory elements or (iv) had high GC-content (see Methods). In brief, these had a median heterozygosity rate of 71%, 97% had motif sizes between 2-6 bp, 8% overlapped gene coding regions or UTRs and 46% overlapped annotated regulatory elements. Descriptive statistics of this set of TRs are shown in Supplementary Figure 1 and Supplementary Table 1.

We generated genotypes for these TRs in a set of 168,554 unrelated individuals of European ancestry from the UKB using ExpansionHunter^8^ analysis of 150bp paired-end Illumina genome sequencing (GS) reads. To ensure that we retained only TRs with high quality genotypes, we also genotyped this same set of TRs in 1,027 individuals from the All of Us (AoU) cohort that had both Illumina GS and Pacific Biosciences (PacBio) HiFi GS. Using genotypes derived from the long-read GS data as a gold standard, we applied quality filters (see Methods), retaining a final set of 36,085 high-quality polymorphic TRs (Supplementary Table 1) that were used for PheWAS with 30,291 binary, quantitative and categorical traits in the UKB cohort (Supplementary Table 2). Associating average allele size of each TR with every trait, in total we observed 5,378 pairwise TR:trait associations that passed a Bonferroni-corrected significance threshold of p<1.45x10^-10^ (Figure 1) (Supplementary Tables 3 and 4). Genomic inflation was well controlled, with λ=1.061 (Supplementary Figure 2). However, as a result of the LD structure of the human genome that frequently results in multiple variants within a locus co-segregating on a single haplotype, we considered that many of these signals were likely to be indirect associations resulting from linkage between the genotyped TR and other causal variant(s) at the same locus. We therefore utilized two complementary approaches to discern the subset of loci where the TR was the most likely causal variant underlying the observed trait associations, namely statistical fine mapping with CAVIAR and conditional analysis applied to both TR and SNV genotypes.

**Figure 1.**
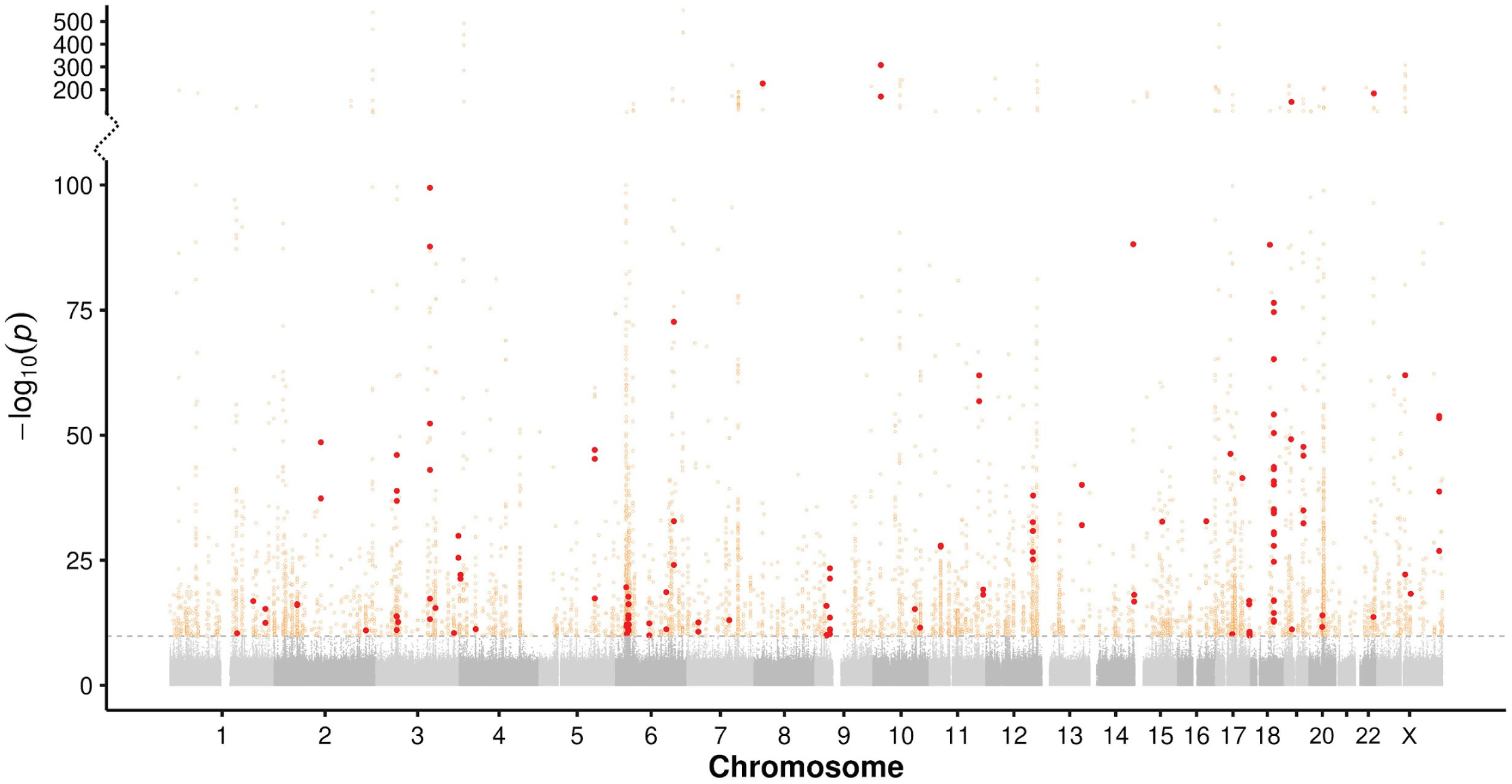
Manhattan plot showing results of PheWAS between 36,085 TRs and 30,291 traits in 168,554 individuals from the UKB. Associations that exceed a Bonferroni-corrected significance threshold of p<1.45x10^-10^ (horizontal dashed line) are shown as colored points. High-confidence causal variants, corresponding to those scored as likely causal by two independent methods, are shown as red filled points, while all other significant associations are shown as open orange circles. Note the discontinuous y-axis and modified scale used to show results with -log10 p >100, which was used to display strong association signals observed at several loci. Full results are shown in Supplementary Tables 3 and 4.

For each of the 5,378 significant TR:trait associations, we first performed association analysis of the same trait with SNVs located within ±500kb of the TR. We then selected the most significant SNV per TR:trait pair and repeated the association test between the TR and trait after dividing the cohort based on genotype at the lead SNV. Doing so identified 864 TR:trait pairs that retained Bonferroni-significance and were considered as putative independent associations by conditional analysis (16% of the original set). For fine mapping analysis with CAVIAR, we analyzed the TR together with the top 100 most significantly associated SNVs per locus. Here, we observed 355 TR:trait pairs where CAVIAR ranked the TR as the most likely causal variant at the locus and which were considered as putative independent associations by fine mapping (6.6% of the original set). Finally, to ensure we identified a stringent set of causal associations, we took the intersection of both of these approaches, defining 101 TR:trait pairs (1.9% of the original set) that we considered as high confidence causal associations scored by two independent methods (Supplementary Table 5).

### Novel causal associations are driven by common polymorphic variation of TRs rather than extreme expansions

The set of high-confidence causal associations included eight TRs that are known to undergo rare expansion to unusually large size, including six that have been shown to cause dominant repeat expansion disorders (REDs) and two others (*BCL2L11* and *CBL*) of indeterminate pathogenicity^13,25^. For four of these six REDs, the associated traits we identified by PheWAS were consistent with the known phenotypic effects of extreme expansions (Supplementary Figure 3), thus indicating that our analysis pipeline correctly identified true causal relationships with TR length: “Huntington’s disease” associated with increased length of a coding CAG repeat in *HTT* (p=7.9x10^-23^)^26^, “Death due to motor neuron disease” associated with a GGCCCC repeat intronic within *C9orf72* (p=1.4x10^-16^)^27,28^, “Hereditary corneal dystrophies” associated with a CAG repeat intronic in *TCF4* (p=6.3x10^-66^)^29^, and “Myotonic disorders” associated with a CAG repeat in the 3’UTR of *DMPK* (p=2.1x10^-48^)^30^.

In order to test whether the other causal associations we identified were similarly attributable to rare expanded TR alleles, we repeated the association test after removing individuals who carried alleles in the top 5% tail of the allelic distribution at each TR. Results of this analysis (Figure 2, Supplementary Table 6) showed that while signals for these four known RED loci were abolished, as expected, nearly all other causal associations we detected by PheWAS retained significance, thus indicating that all other TR associations we detected are attributable to common allelic variation rather than rare expanded alleles. This includes novel associations between length of the *TCF4* repeat with impedance of arm, leukocyte count and lymphocyte count. Variations in these traits have not been reported in patients with the RED Fuchs endothelial corneal dystrophy, and which all therefore appear to be attributable to common allelic variation of this TR. Similarly, for a polyglutamine TR in the *AR* gene, although rare expansions of this TR cause spinal and bulbar muscular atrophy, the associations we observed between polymorphic CAG repeat alleles with “Balding pattern” in males are consistent with prior studies that have demonstrated a role for polymorphism of this TR in male hair loss^31^. Three other TRs in our causal set of associations (a poly(GCC) motif in the 5’UTR of *AFF2,* a poly(CGC) motif in the 5’UTR of *BCL2L11* and a poly(CGG) motif in the 5’UTR of *CBL*) are all known to undergo rare expansion causing allelic methylation, transcriptional silencing accompanied by folate-sensitive fragile sites^25,32^. In the case of *AFF2*, these expansions also cause the FRAXE RED^33^. However, our analysis shows that, as with *TCF4*, the associations of length variation in these TRs with blood cell traits are not the result of extreme expansions, but instead that common polymorphic variation underlies these phenotypic effects (Supplementary Figure 4).

**Figure 2.**
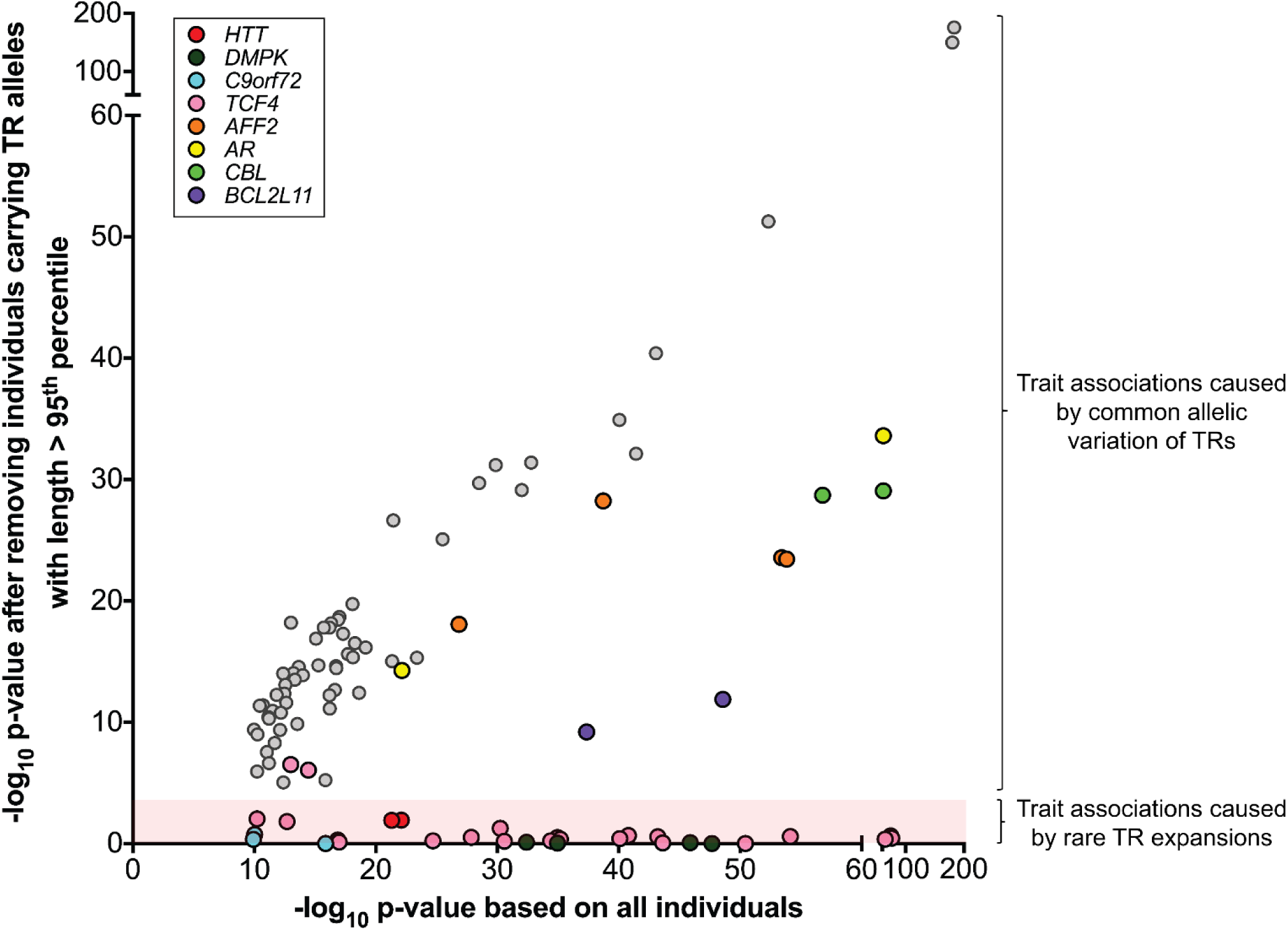
Novel TR associations are driven by common allelic variation rather than rare expansions. We repeated association analysis for each high confidence causal TR:trait pair after excluding any individual carrying an TR allele in the top 5% of the allelic distribution. After excluding long alleles, association signals that matched the known pathogenic effects of four known REDs disappeared (red shaded region). In contrast, nearly all other causal associations we identified remained significant, indicating that these are attributable to common allelic variation rather than rare highly expanded alleles. Colored points indicate eight TRs in the set of high confidence causal associations that are known to undergo rare expansion.

Notable among the novel causal associations is a coding poly(CCA) motif within exon1 of *HRCT1*, a gene of unknown function but which in GTEx data shows highest expression in arterial tissue (Supplementary Figure 5)^34^. This TR is extremely polymorphic and encodes a histidine-rich amino acid tract that showed a strong negative causal association with incidence of high blood pressure (p=4.1x10^-24^) and use of blood pressure medications (p=2.9x10^-14^) (Figure 3). The risk imparted by this TR is such that individuals carrying the shortest 5% of *HRCT1* TR alleles have, on average, an 11% higher risk of hypertension than those carrying the longest 5% of alleles.

**Figure 3.**
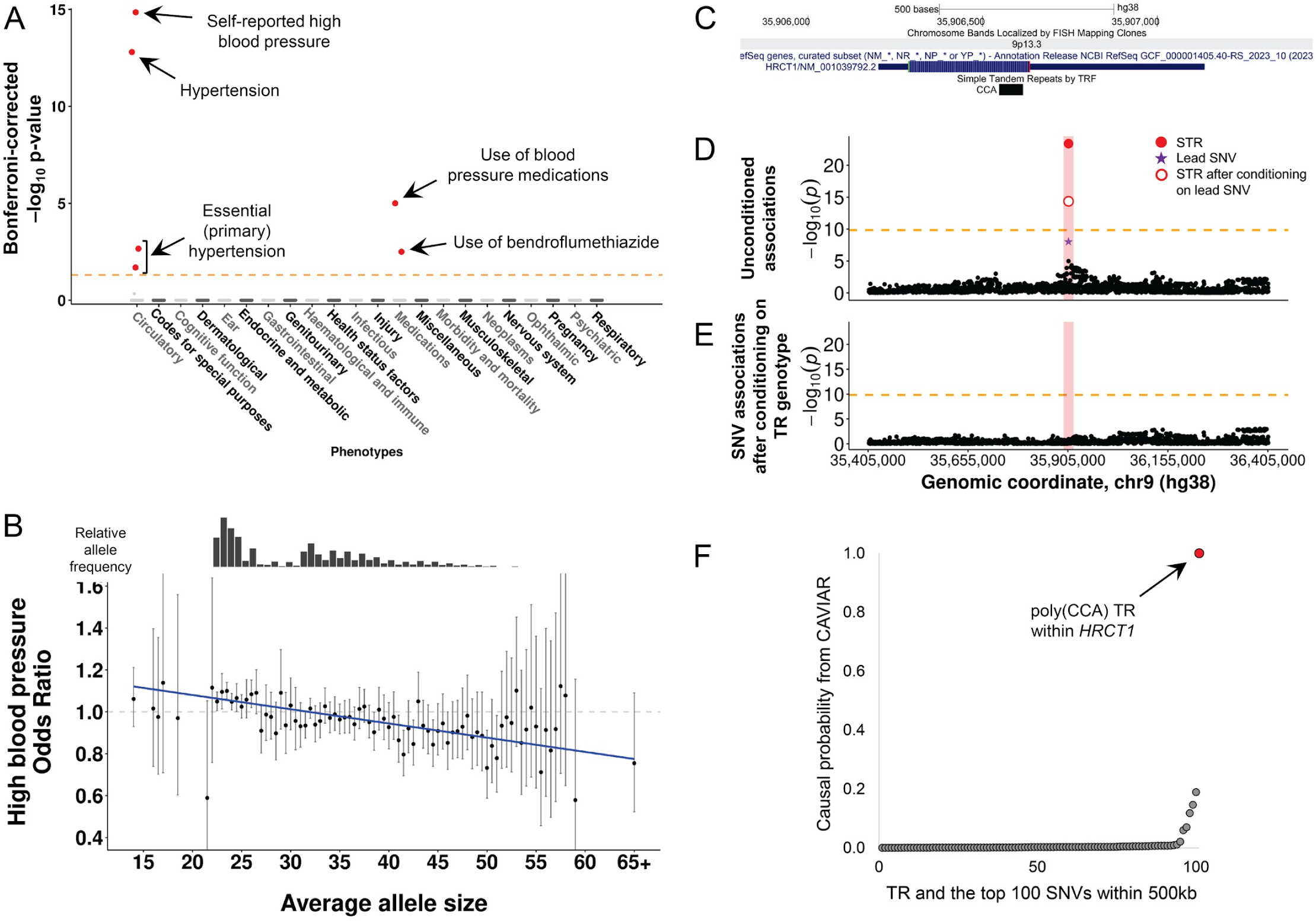
A highly polymorphic coding CCA repeat in *HRCT1* is causally associated with risk of hypertension. **(A)** Results of PheWAS for the TR in *HRCT1*. Traits are grouped into physiological categories (x-axis) plotted against the -log10 Bonferroni-corrected p-value per trait (y-axis). Significant causal associations are individually labeled and shown as filled red points to indicate negative directionality of effect. **(B)** Relative risk of high blood pressure versus average length of the *HRCT1* CCA repeat. Odds ratio per allele is shown by the black dot with vertical lines representing the 95% confidence intervals. To ensure robust estimates of odds ratios, we only plot data for alleles with ≥50 individuals. The bar plot above shows the relative frequency of averaged TR alleles in the UKB cohort. **(C)** Screenshot from the UCSC Genome Browser showing the location of the poly(CCA) motif within *HRCT1*. **(D)** Length of the *HRCT1* CCA repeat (red dot) is the most strongly associated variant in the region with high blood pressure. **(E)** Results after conditioning the same SNVs as shown in (D) based on average genotype of the *HRCT1* repeat. The horizontal dashed line indicates the Bonferroni significance threshold of p<1.45x10^-10^. **(F)** Fine mapping analysis of variants in the *HRCT1* locus with CAVIAR ranks the poly(CCA) motif within *HRCT1* (red dot) as the most likely causal variant underlying risk of high blood pressure.

We also observed novel causal TRs that overlap genes with prior evidence implicating them with the same trait found by PheWAS, providing circumstantial evidence to support a functional role for these TRs. For example, we identified a poly(CGC) motif within the 5’UTR of *GNB2* that was causally associated with pulse rate (p=9.6x10^-14^) (Figure 4). We found that individuals carrying the shortest 5% of *GNB2* TR alleles have, on average, a pulse rate that is 0.86 beats per minute lower than those carrying the longest 5% of alleles. In agreement with this, a prior study identified missense mutations in *GNB2* in sick sinus syndrome 4, a disorder characterized by atrioventricular conduction defects^35^. Similarly, we found a poly(AC) motif in the 3’UTR of *WNT9A* that was causally associated with standing height (p=5.2x10^-16^) (Figure 4). Here, individuals who carry the shortest 15% of *WNT9A* TR alleles are an average of 3.8mm taller than those who carry the longest 15% of alleles, in agreement with multiple studies in model systems that have shown a key role for *WNT9A* in regulating synovial joint formation and chondrocyte differentiation^36,37^.

**Figure 4.**
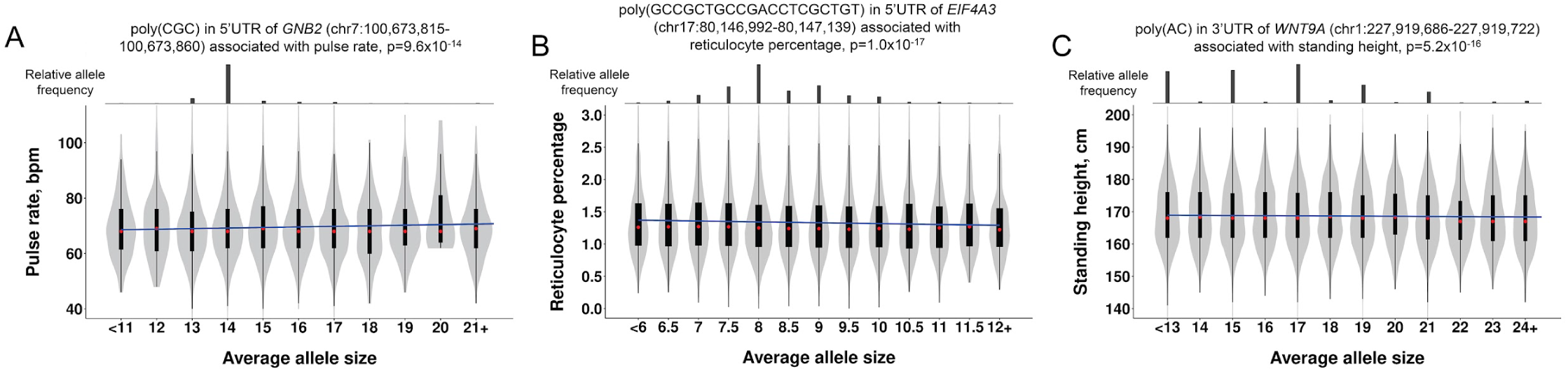
Causal TR associations underlying quantitative traits in the UKB. **(A)** A CGC repeat within the 5’UTR of *GNB2* associated with pulse rate. **(B)** A 20mer repeat within the 5’UTR of *EIF4A3* associates with reticulocyte percentage. **(C)** An AC repeat within the 3’UTR of *WNT9A* associates with standing height. Bar charts above each plot indicate the relative frequency of averaged TR allele sizes in the UKB cohort. The regression slope is shown as a solid blue line and we show the trait distributions per allele size. Within each violin, red circles show the medians; box limits indicate the 25^th^ and 75^th^ percentiles; whiskers extend 1.5 times the interquartile range from the 25^th^ and 75^th^ percentiles.

In order to provide further evidence of the causality of TRs in our high confidence set, we performed conditioning of local SNV associations based on the TR genotype. In addition to results for *HRCT1* (Figure 3), other example plots before and after this conditional analysis are shown in Supplementary Figure 6. For 57 of the 101 high-confident causal associations, we observed that the TR was the most strongly associated variant in the region. In addition, most loci also showed multiple SNVs that were significantly associated with the same trait. However, after conditioning these SNVs based on the genotype of the causal TR, in nearly every case, all SNV associations in the region were nullified, thus providing further evidence that the TRs we identified represent the true causal variants at these loci.

### Replication analysis in the All of Us cohort

We next sought to replicate the set of high-confidence causal associations identified in our discovery PheWAS using the All of Us cohort^38^. We identified phenotypes available for All of Us participants that matched those in UKB and had sufficient sample size for replication analysis for 31 of the 73 causal associated traits. We utilized TR genotypes from GS data from 88,406 All of Us individuals, comprising 58% European ancestry, 25% African ancestry and 17% Latino/Native American ancestry. Despite having an average available sample size in All of Us that was only 40% of that in the UKB, of the 31 causal associations tested, 23 (74%) successfully replicated with p<0.05 and the same direction of effect as observed in the UKB discovery cohort (Figure 5, Supplementary Table 7).

**Figure 5.**
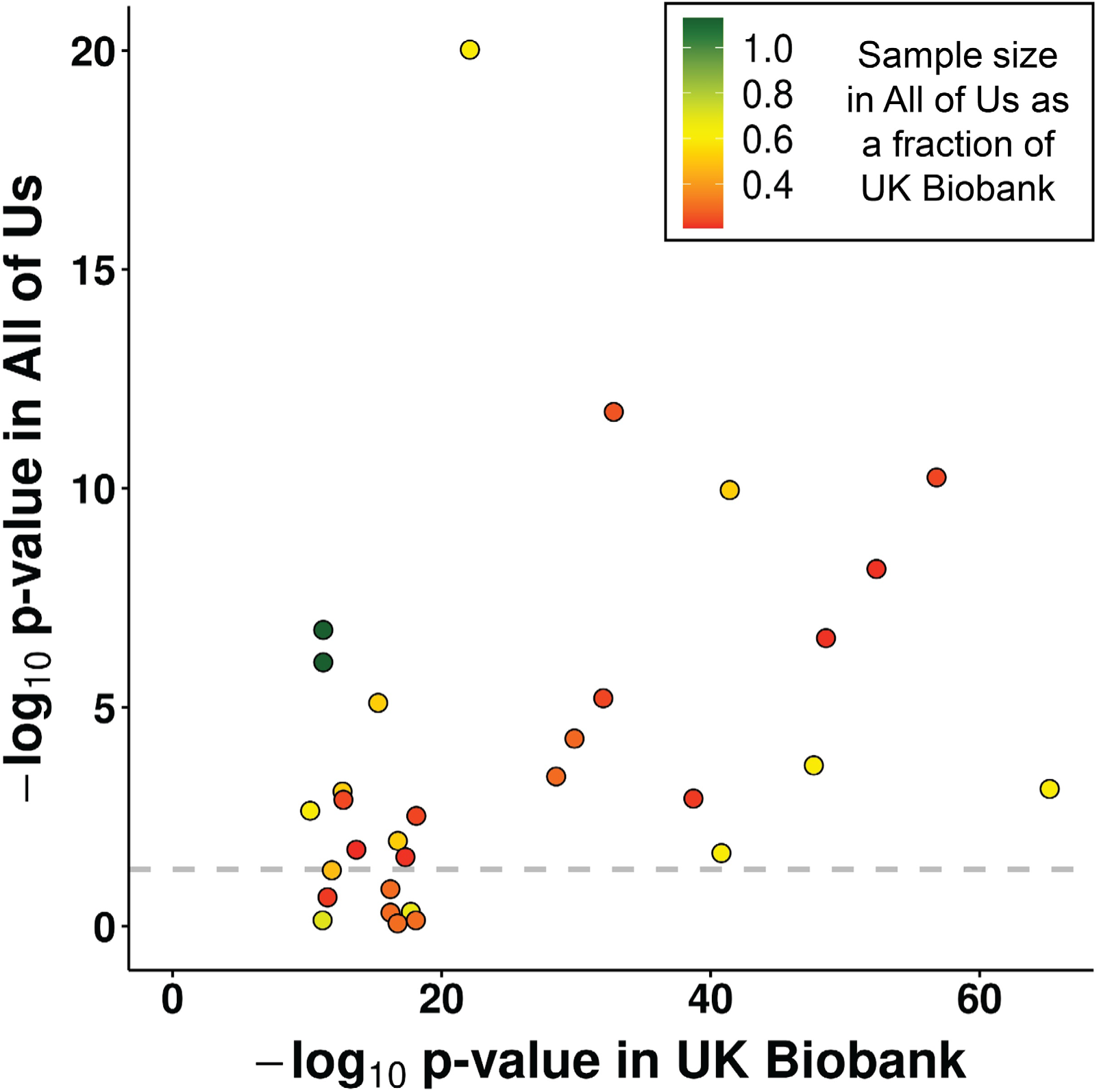
Replication analysis in the All of Us cohort. Using data from the All of Us cohort, we performed replication analysis for 31 high-confidence causal associations identified in the UKB discovery cohort. The dashed horizontal line indicates p=0.05 in the All of Us cohort while the color of each point indicates the relative sample size per trait in the All of Us replication cohort compared to the UKB discovery cohort. Full results are shown in Supplementary Table 7.

### Causal TRs are strongly enriched for functional effects on gene expression and epigenetics

To assess the functional effects of TR variants on local gene expression and epigenetics, we genotyped the set of TRs used for PheWAS from available GS data and compared these with DNA methylation and RNAseq data for 49 tissues generated by the GTEx project^39,40^, identifying TRs that act as expression or methylation QTLs (eQTLs and mQTLs) (Supplementary Tables 8 and 9). Considering all TRs that showed significant trait associations by PheWAS in the UKB, these were 4.9-fold enriched versus the null for eQTLs (p=1.5x10^-193^, two-tailed Fisher’s exact test) and 3.5-fold enriched for mQTLs (p=6x10^-103^). Considering the subset of TRs that were deemed as high-confidence causal variants for trait associations, these were 11.2-fold enriched for eQTLs (p=9.2x10^-12^) and 4.7-fold enriched for mQTLs (p=1.2x10^-5^) (Figure 6).

**Figure 6.**
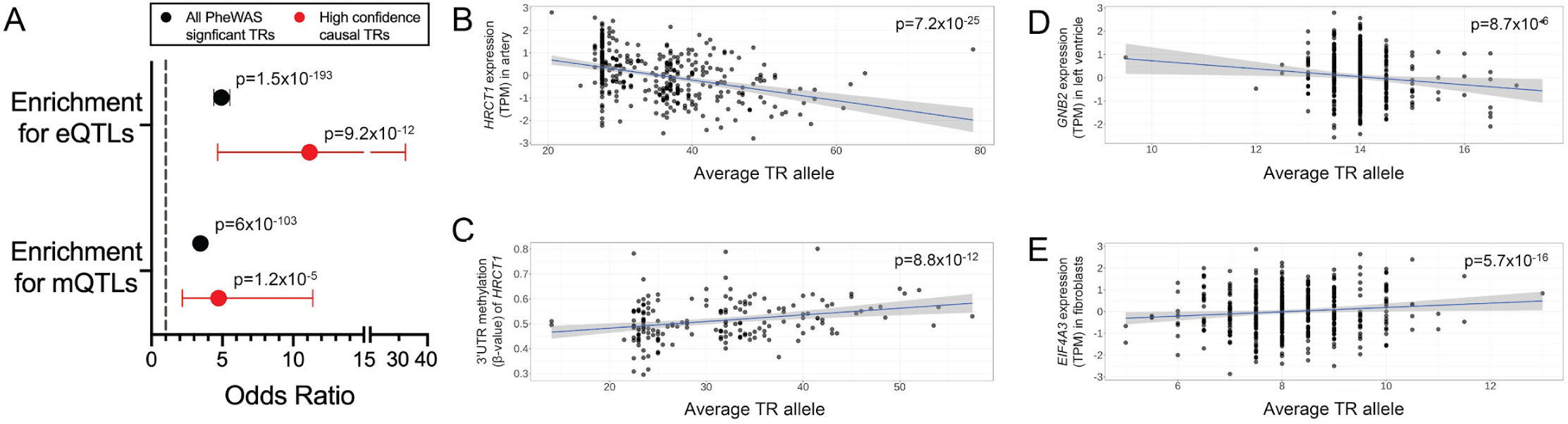
Length variation of causal TRs associates with local gene expression and DNA methylation. **(A)** Causal TRs showed significant enrichments for both eQTLs and mQTLs in the GTEx cohort and were often associated with the same gene as which the TR was located within. Horizontal lines indicate 95% confidence intervals of enrichments. **(B and C)** Results for an exonic poly(CCA) motif in *HRCT1* associated with both normalized *HRCT1* expression (shown for aorta tissue) and DNA methylation of a CpG (cg21518683) in the 3’UTR of *HRCT1* (shown for transverse colon tissue). **(D)** a poly(CGC) motif within the 5’UTR of *GNB2* associated with *GNB2* expression (shown for left ventricle tissue). **(E)** a poly(GCCGCTGCCGACCTCGCTGT) motif within the 5’UTR of *EIF4A3* associated with *EIF4A3* expression (shown for cultured fibroblasts).

For example, the coding poly(CCA) repeat in *HRCT1* that is associated with risk of hypertension showed both a negative association with *HRCT1* expression and a positive association with methylation level of a CpG in the 3’UTR of *HRCT1* (Figure 6). Multiple other causal TRs were also associated with expression level of the overlapping gene and/or other genes *in cis*, suggesting that the mechanism of action of many of the TR associations we identify is through altered gene expression and/or epigenetics.

## Discussion

Here, we performed the first phenome-wide analysis based on direct genotyping of the most polymorphic and unstable TRs in the genome, identifying many novel causal phenotype associations linked with length variation in these TRs that have been overlooked in prior SNV-based GWAS. In support of our results, our analysis identified both known causal effects associated with several recurrent REDs and also replicated effects of common TR variation at *CBL* and *BCL2L11* that were reported in a recent study that utilized imputation of TR alleles^24^.

Of note, our analysis identified causal effects on traits linked with common allelic variation at three TRs (*TCF4*, *AR* and *AFF2*), while rare expansions of these same loci are known to underlie REDs which manifest with completely different phenotypes. Thus, some TRs can exert divergent effects at different size ranges, with variation in shorter polymorphic alleles acting to modify common traits while rare extreme expansions of the same TR cause REDs with quite different phenotypic features.

Fine mapping of TR variants is challenging^24^. As a result, it is possible that our definition of high confidence causal TRs based on the intersection of two approaches is overly-conservative, and it is likely that some TRs that have a true causal influence on human traits do not meet these stringent criteria. For example, conditional analysis may yield false negatives when multiple causal variants occur within a locus, while fine mapping approaches can have limited ability to resolve a single causal variant in regions of extended LD. Indeed, several functionally compelling candidates were identified by fine mapping as the most likely causal variants underlying the observed associations, but which did not meet Bonferroni significance upon conditional analysis, *e.g.* a poly(GCGG) motif in the 5’UTR of *CHRNA3*, a nicotinic acetylcholine receptor, that associated with pack years of smoking. Future work using larger sample sizes and incorporating diverse ancestries will yield improved power to resolve additional effects of TR variants.

Our study has several potential strengths and limitations: (i) In comparison to previous studies that have relied upon imputation-base approaches to statistically infer TR genotypes^21,24,41^, we performed TR genotyping directly from GS reads and thus our study likely had improved genotyping of highly mutable loci and rare TR alleles that typically have low imputation accuracy. (ii) We tested a limited set of TRs with motif sizes between 2-20 bp, but which are highly enriched for the most polymorphic and unstable TRs in the genome and which preferentially overlap functional and regulatory elements. (iii) We utilized average TR allele size in our association model and thus our analysis had reduced power to identify more complex non-linear relationships and did not consider sequence variations within TRs. Despite this, we still identified associations with several known REDs, where disease associations exhibit a threshold effect^13^. (iv) Due to a limited sampling of diverse ancestries in the UKB, our analysis only utilized individuals of European ancestry and thus missed associations that may be specific to non-European populations. (v) By using ExpansionHunter, an algorithm that can provide estimates of TR alleles that exceed the read length of GS, we were able to identify associations with TRs that exhibit very long alleles, such as those at *HRCT1, EIF4A3* and at several RED loci where causal alleles are typically >150bp in size.

Our study highlights the contribution of multi-allelic TRs as a potential contributor to the “missing heritability” of SNV-based GWAS, emphasizing the importance of considering TRs in fine mapping studies.

## Materials and Methods

### Genotyping of tandem repeats in the UK Biobank (UKB) and All of Us (AoU) cohorts

We derived a catalog of TRs with motif sizes ranging from 2-20bp that we hypothesized would be enriched for functional effects and which are either highly polymorphic or were observed to undergo rare expansion in the human population using the following approach:

1. We utilized both a catalog of ∼174,000 TRs with motif sizes ranging from 2-20bp genotyped with ExpansionHunter (v5.0.0)^8^, and a catalog of 798,697 TRs with motif sizes ranging from 2-6bp genotyped with hipSTR (v0.7)^10^. Using these two tools, we performed genotyping of high coverage PCR-free Illumina GS data for 2,504 individuals from the 1000 Genomes Project^42^. Based on the resulting genotypes, we then extracted any TR which showed either (i) ≥20 different alleles with either genotyping tool, or (ii) ≥5 different alleles with either genotyping tool for those TRs that overlapped candidate *cis*-regulatory elements defined by ENCODE (“ENCODE cCREs” track from the UCSC genome browser), regulatory regions defined by GeneHancer (“Enhancers and promoters from GeneHancer (Double Elite)” track from the UCSC genome browser), or genic regions defined as either exon, 5’UTR, 3’UTR, upstream, downstream or splicing by ANNOVAR^43^ based on Ensembl gene annotations.
2. Based on TR annotations of the hg38 assembly using TRFinder^44^ (“Simple repeats” track from the UCSC genome browser), we selected any TR with motif size between 2-10bp that was composed purely of C and G bases.
3. We utilized both ExpansionHunter Denovo (v0.9.0) ^9^ and STRetch (v0.4.0) ^11^ to screen a set of 22,579 individuals of diverse ancestry with high coverage PCR-free Illumina GS data, comprising (i) 10,961 individuals from the TOPMed Women’s Health Initiative cohort, (ii) 9,114 individuals from the TOPMed BioMe cohort, and (iii) 2,504 individuals from the 1000 Genomes Project. In each of these three cohorts, we identified TRs showing long outlier allele sizes. We defined these as any TR with ≥3 in-repeat reads in at least one individual as assessed by ExpansionHunter Denovo, or from the output of STRetch, any TR that passed all of the following criteria in at least one individual in any of the three cohorts: FDR q< 0.1, locus coverage >0, allele size ≥15, allele size >99.5^th^ percentile of the cohort for that TR, allele size ≥5 above the cohort median for that TR.

From these, we selected those TRs that had a span in the hg38 reference genome of ≤150bp, resulting in a set of 48,913 TRs located on chr1-22 and chrX that we retained for further analysis (Supplementary Table 1).

Prior to association analysis, we performed quality filtering of genotypes generated by ExpansionHunter for these TRs using data generated by the All of Us project. All of Us has released data from both short-read GS with Illumina and long-read GS with PacBio HiFi technology in 1,027 individuals. We performed genotyping of the set of 48,917 TRs in both the Illumina data using ExpansionHunter and in the PacBio data using TRGT (v0.4.0) ^45^, with identical TR definitions used for both tools. We first quality filtered the TRGT genotypes, removing individual TR genotypes supported by only a single spanning read and those with purity scores <0.75. After applying these quality filters, we then retained only those TRGT genotypes in each sample where both alleles remained. First, we removed 439 TRs that showed no variation in the genotypes output by TRGT. Then, for each individual and TR, we calculated average TR genotypes output by both ExpansionHunter and TRGT. For each TR we performed Spearman correlation between genotypes generated by the two different sequencing technologies. For those TRs with median purity scores from TRGT ≥0.75, we removed 5,076 TRs that showed Spearman R<0.5 between the ExpansionHunter and TRGT genotypes. In addition, we removed 7,313 TRs that showed low variation in the genotypes from ExpansionHunter profiling of UKB samples, defined here as standard deviation <0.5. After these filters, we used a final set of 36,085 high-quality, highly polymorphic TRs that are enriched in genic and regulatory regions for PheWAS (Supplementary Table 1).

### PheWAS analysis in the UK Biobank

Collection of the UKB data was approved by the Research Ethics Committee of the UKB obtained under application 32568. All study participants provided informed consent, and the protocols for UKB are overseen by The UKB Ethics Advisory Committee, see https://www.ukbiobank.ac.uk/ethics/. From the set of 200,000 individuals with Illumina 150 bp paired-end whole GS data, we defined 188,915 individuals of European ancestry using an approach based on principal component analysis of a set of ∼64,000 high quality LD pruned SNVs with minor allele frequency (MAF) >5%, as described previously^22^. In brief, GCTA (v1.93) was used to calculate the first 40 Principal Components (PCs), which were used to predict the ancestry of each individual. Since only a small minority of individuals recruited to the UKB cohort are of non-European ancestry and are, therefore, highly under-powered in PheWAS, we focused our analysis on those individuals of European origin. We first selected all samples predicted as European ancestry and recalculated PCs without use of a reference population using one third of individuals, before projecting the remaining two thirds of individuals onto this. We removed outlier samples using within group PCA. In addition, we kept only those samples with self-reported ancestry as “White”, “British”,“Irish”,“Not listed in table”, “Any other white background”, “Other ethnic group”, or “Prefer not to answer”. Further sample-level filters were applied as follows:

1. Using pairwise kinship coefficients provided by the UKB, for samples with 2^nd^ degree relationships or higher (kinship coefficient >0.0883) we retained only a single unrelated individual.
2. We removed 199 individuals with predicted sex chromosome aneuploidy.
3. We removed 241 individuals who we were notified had withdrawn their consent for inclusion in the UKB study.

After these quality control (QC) steps, we retained data for 168,554 unrelated individuals of European ancestry. Using the UKB DNAnexus Research Analysis Platform, we performed genotyping of 36,085 TRs from the GS data with ExpansionHunter (v5) (Supplementary Table 1).

In order to identify traits associated with the presence of TRs, we utilized the mean TR genotype present in each sample at each locus in association analysis. We utilized phenotype data for individuals in the UKB derived from a total of 30,291 ICD10 codes and quantitative, categorical and binary traits (Supplementary Table 2), accessed through UKB application number 82094. Before performing association testing, phenotype data were processed as follows:

1. For phenotypes with multiple categories, *e.g.* ICD10 codes, we considered each category as a separate binary trait. If a subject had multiple entries for the same category, they were assigned as “1” (*i.e.* positive for that trait) if any of the instances were positive. We removed from analysis any categories labeled as “Prefer not to answer”, “Unsure” or “Do not know”.
2. ICD9 and ICD10 codes were merged into one using General Equivalence Mapping (GEM) files downloaded from https://www.cms.gov/Medicare/Coding/ICD10. We restricted controls to subjects who did not have a positive diagnosis for that phenotype.
3. In addition to using each individual ICD code as a separate trait, ICD codes were also grouped into higher level codes to yield additional summary traits with increased sample size. For example, the ICD10 code for Schizophrenia is F20, which is composed of 10 different sub-codes (*e.g.* F20.0 “Paranoid schizophrenia:, F20.2 “Catatonic schizophrenia”, F20.9 “Schizophrenia, unspecified”, *etc.*), with each subcategory containing between 4 and 894 individuals. Here, we created a new summary code for “Schizophrenia_group” comprising all sub-categories under F20 which included 1,485 individuals who were positive for any subtype of schizophrenia. Similar groupings were performed for any higher level ICD code that comprised multiple subtypes.
4. For phenotypes with >2 categorical outcomes, each outcome was assigned an integer value and these were analyzed as quantitative traits. *e.g.* the phenotype “Alcohol intake frequency” included six categories: “Daily or almost daily”, “Three or four times a week”, “Once or twice a week”, “One to three times a month”, “Special occasions only” and “Never”. Here, each sample was assigned an integer value ranging from 1 to 6, with each value corresponding to the six ordered frequencies for this phenotype. Categorical traits with just two separate categories were considered as binary traits.
5. For quantitative traits with integer values, where a subject had multiple instances, we utilized the mean of all instances rounded to nearest integer
6. For quantitative traits with continuous values, where a subject had multiple instances, we utilized the mean of all instances.

For the 1,349 quantitative traits analyzed, we applied a rank based inverse normal transformation and required a minimum sample size of 50 phenotyped individuals and standard deviation ≥0.001 to be included in association analysis. For the 28,942 binary traits analyzed, we required a minimum sample size of at least 10 individuals with the trait to be included in the analysis.

Association analysis was performed using *REGENIE* (v3.1.3) ^46^, incorporating covariates of sex, age, age squared, GS insert size and the top five PCs derived from analysis of SNVs to account for ancestry. For binary traits, we utilized the Saddle Point Approximation function to reduce the type I error rate. To avoid possible technical effects on TR genotypes, associations were performed separately in samples sequenced by deCODE and Sanger Center, before combining results using z-score based meta-analyses in *METAL* (PMID: 20616382). To ensure that we only reported robust signals, we required that associations showed the same direction of effect and with p<0.05 in both sub-cohorts. For loci on the X chromosome, samples were separated by both sequencing center and sex and associations performed separately in the four resulting sub-groups (sequencing center + sex) before combining results using *METAL*, requiring that at least two of the four sub-groups met the minimum sample size requirements as stated above, at least two of the four sub-groups yielded p<0.05 and all showed the same direction of effect.

We applied multiple testing corrections using both a Bonferroni approach based on the number of TRs and independent traits analyzed. However, given that many phenotypes in the UKB are highly correlated (*e.g.* neutrophil count and neutrophil percentage), it should be noted that multiple testing corrections tend to be overly stringent. While some approaches remove correlated phenotypes and *e.g*. retain only a single trait with the largest sample size, we chose not to pursue this method, as it will often lead to missing the primary trait associated with a variant. However, to estimate a better calibrated Bonferroni threshold in our analysis, we calculated the number of nominally independent traits tested by performing a pairwise correlation between all traits utilized in our PheWAS. Then, considering a threshold of R<0.5 between any two traits as indicative of independence, we calculated the minimal set of independent traits(n=9,531) used in our analysis. Based on this and considering the n=36,085 TR genotypes that were used in PheWAS, we utilized an adjusted Bonferroni correction threshold in our meta-analysis of p<1.45x10^-10^ (p=0.05 / 9,531 / 36,085).

For creating plots of grouped PheWAS results, we assigned each trait reported by the UKB into one of 22 different categories based on shared physiological systems, tests or treatments. To create these groupings, phenotype codes were mapped to ICD10 chapters as listed in Supplementary Table 1 of Wang *et al*.^47^. Phenotypes that did not have a category assigned with these annotations were further annotated using categories provided within the UKB Data Showcase. Finally, we applied manual curation, removing categories that were composed of a very small number of traits and making reassignments to improve the consistency of groupings.

### Identification of high confidence causal TRs using conditional analysis and fine mapping

In order to assess whether TRs were independently associated with a trait, we performed two separate analyses to determine a set of high confidence causal variant(s) at each locus detected in our PheWAS.

In the first of these, we performed conditional analysis by separating individuals by genotype state based on the lead associated SNV with the trait in question for each region. We utilized SNV genotypes derived from GS data from UKB participants. We removed SNVs that had either minor allele count<100, missingness rate >5%, HWE<10^-300^, quality score (QUAL) <30, mapping quality (MQ) <40, read depth (DP) <10, genotype quality (GQ) <20, strand bias (SB) <0.25 or >0.75 and allelic balance for heterozygous calls (ABhet) <0.2. We extracted all remaining SNVs within ±500kb of each TR. For each TR:trait pair that was significant in PheWAS, we performed association analysis between that trait and all SNVs located within ±500kb using REGENIE. This was performed separately in each of the two sequencing sub-cohorts, before combining p-values together using METAL. We considered TRs as potentially independently associated as those that retained a Bonferroni-corrected association with the trait of p<0.05 after conditional analysis based on the minimal set of 9,531 independent phenotypes used (nominal p<5.24x10^-6^).

For conditional analysis of each TR based on local SNVs, for each TR:trait pair we selected the most significant SNV with MAF >0.01 and then divided individuals into three groups based on their genotype at this lead SNV (*i.e.* AA, AB and BB), which were further divided based on the two sequencing sub-cohorts. In each of these six groups (genotype at lead SNV x sequencing cohort), we required a minimum of 50 genotyped individuals with the trait for quantitative traits and 10 genotyped individuals with the trait for binary traits, utilized REGENIE to repeat the association test with the TR:trait pair, and then merged the resulting p-values using METAL.

We also performed statistical fine mapping using CAVIAR^48^ on the Sanger Center sub-cohort using the top 100 most significantly associated SNVs per locus, as defined above. We calculated LD between SNVs and TRs by pairwise correlation. P-values from REGENIE were converted into z-scores and CAVIAR run using parameters ρ=0.95, γ=0.01 and the maximum number of causal variants was set to 2.

We took a conservative approach to define putatively causal TRs based on the results of both conditional analysis and fine mapping. High confidence causal TRs were considered as those that were both the top ranked variant by CAVIAR and which also retained a Bonferroni-corrected association with the trait of p<0.05 after conditional analysis based on the minimal set of 9,531 independent phenotypes used (nominal p<5.24x10^-6^).

### Conditional analysis of local SNVs based on TR genotypes

To gain additional evidence for causality of TR associations, we performed conditional analysis of SNVs based on TR genotypes at each locus containing a high confidence causal TR. Here, we divided individuals into groups based on their average TR genotype, which were further divided based on the two sequencing sub-cohorts. In each group we removed SNVs with MAF<0.01, missingness >5% or Hardy-Weinberg Equilibrium p<1^-300^ and required a minimum of 50 genotyped individuals with the trait. For each remaining SNV, in each group (genotype at causal TR x sequencing cohort), we utilized REGENIE to repeat the association test of the SNV with the trait, and then merged the resulting p-values across all groups per SNV using METAL.

### Assessing whether association signals result from common allelic TR variation or rare expansion alleles

In order to assess whether causal associations were driven by common variation in TR length rather than rare expansions or contractions, for each causal variant we repeated association analysis after excluding individuals who carried a TR allele that lay in the upper tail of the allelic distribution at that locus. Allele length percentiles were calculated separately per TR based on diploid genotypes in each of the two sequencing sub-cohorts. For each TR, we excluded any individual who carried an allele that was >95^th^ percentile and then repeated association analysis using the same method as described above.

### Replication analysis in All of Us

We curated data for 27,660 traits for 245,394 individuals with Illumina GS data in the All of Us (AoU) v7 release. These traits were derived from a total of 26,329 unique OMOP codes (v5.3.1) corresponding to binary, quantitative, and categorical data. For ∼94% (26,134/27,660) of these traits, each mapped to a single OMOP code. The remaining 1,526 traits were derived from 195 duplicated OMOP codes (194 from the AoU domain “Survey Questions” and 1 from the domain “Labs and Measurements”). Post-review and processing, all OMOP codes corresponding to categorical data were converted into either binary or quantitative traits depending on the nature of the underlying data. Prior to association testing, phenotype data were processed as follows:

1. The underlying data in the AoU domains “Conditions”, “Procedures”, “Drug Exposures”, and “Devices” are by nature binary, so all data generated from these domains were classified as binary traits. An individual was assigned “1” for a trait if they were positive for it (*i.e.* had the condition, were exposed to the drug, etc.), and “0” if not.
2. The underlying data in the AoU domains “Labs and Measurements”, “Program Physical Measurements” and “Survey Questions” were treated as either quantitative or categorical traits.
3. Those traits with multiple quantitative values were treated as quantitative traits. Where an individual had multiple values recorded, presumably representing repeat measurements taken at different times, we utilized the mean of all values. For the majority of quantitative traits, individuals had data recorded in multiple different units of measure. For example, height was recorded as either centimeters or inches in different individuals. For each such trait, we only retained data for those individuals that were recorded in the unit with the greatest number of unique individuals, excluding all cases where units were listed as “No matching concept”, “No value”, and “NA”.

We observed that some traits had values that were not physiologically possible, suggesting the data were erroneous. For example, body temperature was listed as either 0 or 100 degrees Celsius in some individuals. We therefore performed quality filtering to remove such values, as follows:

1. For traits where the unit was “percent”, we set lower and upper limits of 0 and 100, removing any values outside this range.
2. For traits recorded in other units, we identified 167 that had at least one individual with a value >10 standard deviations from the mean. For these traits, we manually set reasonable lower and upper limits for each trait to ensure that all values remained within physiologically possible ranges based on published medical literature, removing any data points that lay outside these ranges.

For categorical data with two possible outcomes (*e.g*., lab results that could be either positive or negative), we considered these as binary traits. Where an individual had multiple values recorded, we considered them positive for that trait if any of the values were positive. For categorical data with more than two possible outcomes after removing those such as “Prefer not to answer”, “Not sure”, *etc*., these were converted to quantitative traits, Here, each outcome was assigned an integer value in ascending order after sorting the possible outcomes. *e.g.,* the OMOP code “1586201, Alcohol: Drink Frequency Past Year” included five categories: “Never”, “Monthly or less”, “2 to 4 per month”, “2 to 3 per week”, and “4 or more per week”.

Here, each individual was assigned an integer value ranging from 1 to 5, with values corresponding to the five ordered frequencies for this trait.

Using tables provided by the UKB (data field 20142) we mapped the set of traits associated with high confidence causal TRs identified in the discovery PheWAS to matching codes provided by All of Us. In addition to this automated matching, we also performed text searching based on keywords and synonyms followed by manual curation. In order to ensure sufficient statistical power for replication, we only retained matching traits where the sample size in the set of genotyped All of Us samples with that trait was ≥20% that available in the UKB cohort. This resulted in a final set of 31 matching TR:trait pairs, corresponding to 20 unique traits, available in All of Us for replication (Supplementary Table 6).

For each of the high confidence causal TRs identified in our discovery PheWAS we performed genotyping of 107,737 individuals from the All of Us v7 data release using Illumina GS data and ExpansionHunter. Where individuals were related, defined by All of Us as those with pairwise kinship scores >0.1, we randomly removed one member of the related pair. We also removed individuals with TR genotyping rate <99%, predicted sex chromosome aneuploidy based on genome-wide read depth analysis using mosdepth^22^ and those with ancestry defined by All of Us as East Asian, South Asian, Middle Eastern or Other. After these filtering steps we retained a total of 88,406 individuals, comprising 51,089 individuals of European ancestry (EUR), 22,248 individuals of African (AFR) ancestry and 15,069 individuals of Latino/Native American ancestry (AMR).

For quantitative traits, where in each case data were available for >20,000 individuals, prior to association analysis, samples were divided into twelve different groups based on both ancestry (EUR, AFR, AMR) and sequencing center/sequencing date (Baylor, University of Washington, Broad Institute prior to September 2019 and Broad Institute post September 2019). In contrast, for binary traits where the number of individuals positive for some traits was <100, to maintain robustness, we performed association analysis using all ancestries and sequencing centers combined as a single group. Associations were performed using *REGENIE*, incorporating the same covariates and options as utilized for the discovery PheWAS in UK Biobank. For quantitative traits, results for each of the 12 sub-groups were combined using z-score based meta-analyses with *METAL*.

### Identification of TR QTLs using GTEx data

To identify associations of TR length with variation in local gene expression and DNA methylation, we utilized GTEx data (release v9) downloaded from the GTEx portal. For the Illumina GS data, samples marked as low quality by GTEx were removed. TR genotypes were generated using ExpansionHunter with the same TR catalog as applied above and converted to average repeat length. QC was performed separately for each sequencing protocol (either with or without the use of PCR during library preparation). We removed outlier samples based on genotype density and PCA plots. Additionally, 68 samples sequenced on the HiSeq 2000 instrument were removed. We removed TRs that did not pass default ExpansionHunter quality filters, had high rates of missing genotypes (>5% for PCR-free samples, >10% for samples sequenced with PCR) or showed low levels of variation in the GTEx cohort (standard deviation <0.5). After these filtering steps, we retained data for 52,855 TRs and 899 individuals, comprising 343 individuals sequenced using a library prepared without PCR amplification and 556 individuals sequenced using a library prepared with PCR amplification.

#### Gene expression data

RNAseq was performed in different tissues using the Illumina TruSeq library construction protocol, pre-processed, aligned and subject to QC as described^49^. Briefly, for each tissue, read counts from each sample were normalized using size factors calculated with DESeq2 and log-transformed with an offset of 1; genes with a log-transformed value >1 in >10% of samples were selected, and the resulting read counts were centered and unit-normalized. The resulting matrix was then hierarchically clustered (based on average and cosine distance), and a chi^2^ p-value was calculated based on Mahalanobis distance. Clusters with ≥60% samples with Bonferroni-corrected p-values <0.05 were marked as outliers, and their samples were excluded. Samples with <10 million mapped reads were removed. For samples with replicates, the replicate with the greatest number of reads was selected.

Gene expression values for all samples from a given tissue were normalized using the following procedure. Genes were selected based on expression thresholds of >0.1 TPM in at least 20% of samples and ≥6 reads in at least 20% of samples. Expression values were normalized between samples using TMM as implemented in edgeR^50^. For each gene, expression values were normalized across samples using an inverse normal transformation. The normalized QC’d matrices and corresponding covariates for the expression data were downloaded from the GTEx Portal. The final expression data contained 49 tissues with between 73 and 706 individuals and 20,000-35,000 genes per tissue.

Association analyses between TR genotypes and expression level of each gene located within ±500kb were performed in two steps using the lm() function in R, incorporating covariates based on the top five principal components from TR genotypes, sequencing protocol (with PCR or PCR-free), sex, age, GS mean insert size and additional PEER factors^51^ provided by GTEx. First, we adjusted the expression data for covariates and extracted the resulting residuals. Second, for any TR:gene pair with ≥60 samples available, these residuals were used as an input for linear regression against average repeat length. We applied a False Discovery Rate correction for multiple testing and considered associations to be significant at 1% FDR (q<0.01).

#### DNA methylation data

DNA methylation at 866,895 CpG positions was assayed in nine different tissues of GTEx individuals with the Illumina MethylationEPIC array. Data were processed and subject to QC as described^40^. Briefly, for each tissue, raw data were processed with ChAMP^52^ (v.2.8.6). We excluded three samples with undetectable or missing methylation values (detection p>0.01) in ≥5% of CpG sites and six samples with a sex mismatch between methylation and DNA sequencing data. For QC, all CpG sites that had detection p>0.01 in more than one sample and that had a bead count <3 in ≥5% samples were excluded. Additionally, cross-reactive CpG sites and CpG sites that either overlapped or had a base-pair extension overlapping common SNVs were filtered out. Finally, CpG sites mapping to sex chromosomes were also removed. After these filtering steps, 754,054 CpG sites passed QC and had their genomic coordinates lifted over from the GRCh37 to GRCh38 human genome build. The QC’d matrices and corresponding covariates for the DNA methylation data were downloaded from the GTEx Portal.

From nine tissues assayed, only the four that had profiles from >60 samples were used for further analysis. From these, we removed outlier samples based on PCA plots. CpG sites that showed low levels of variation (standard deviation of beta values <0.01) within each tissue were removed. The final DNA methylation data comprised four tissues with between 103 and 190 individuals and 540,000-590,000 CpG sites per tissue.

Association analyses was performed between average TR genotypes and DNA methylation level of each CpG site located within ±50kb using the lm() function in R, incorporating covariates based on the top five principal components from TR genotypes, GS protocol (with PCR or without PCR), sex, age, GS mean insert size and additional PEER factors provided by GTEx. First, we adjusted the DNA methylation data for covariates and extracted the resulting residuals. Second, for any TR:CpG site pair with ≥60 samples available, these residuals were used as an input for linear regression against average repeat length. We applied a False Discovery Rate correction for multiple testing and considered associations to be significant at 1% FDR (q<0.01).

## Data Availability

All genotype data generated in the UKB and All of Us cohorts have been returned and will be available through future data releases.
Code utilized for this study is available as follows:
https://github.com/Illumina/ExpansionHunter
https://github.com/bharatij/Global-Ancestry-Assignment
https://github.com/PacificBiosciences/trgt

https://www.internationalgenome.org/data-portal/data-collection/30x-grch38

https://www.ncbi.nlm.nih.gov/projects/gap/cgi-bin/study.cgi?study_id=phs001644.v2.p2

https://biobank.ctsu.ox.ac.uk/crystal/search.cgi

https://www.researchallofus.org/data-tools/workbench/

https://www.ncbi.nlm.nih.gov/geo/query/acc.cgi?acc=GSE213478

https://storage.googleapis.com/gtex_analysis_v8/rna_seq_data/GTEx_Analysis_2017-06-05_v8_RNASeQCv1.1.9_gene_tpm.gct.gz

## Acknowledgements

This research has been conducted using the UK Biobank Resource under Application Number 82094 and the NIH All of Us data under research project “Association studies of tandem repeats”. This work was supported by NIH grants AG075051, NS105781 and HD103782 to A.J.S. and NHLBI BioData Catalyst Fellowship #5120339 to A.M.T.

This work was supported in part through the computational resources and staff expertise provided by Scientific Computing at the Icahn School of Medicine at Mount Sinai and supported by the Clinical and Translational Science Awards (CTSA) grant UL1TR004419 from the National Center for Advancing Translational Sciences. Research reported in this paper was supported by the Office of Research Infrastructure of the National Institutes of Health under award number S10OD026880 and S10OD030463. The content is solely the responsibility of the authors and does not necessarily represent the official views of the National Institutes of Health.

Molecular data for the Trans-Omics in Precision Medicine (TOPMed) program was supported by the National Heart, Lung and Blood Institute (NHLBI). Genome sequencing for “NHLBI TOPMed - NHGRI CCDG: The BioMe Biobank at Mount Sinai” (phs001644.v1.p1) was performed at the McDonnell Genome Institute (3UM1HG008853-01S2). Genome sequencing for “NHLBI TOPMed: Women’s Health Initiative (WHI)” (phs001237.v2.p1) was performed at the Broad Institute Genomics Platform (HHSN268201500014C). Core support including centralized genomic read mapping and genotype calling, along with variant quality metrics and filtering were provided by the TOPMed Informatics Research Center (3R01HL-117626-02S1; contract HHSN268201800002I). Core support including phenotype harmonization, data management, sample-identity QC, and general program coordination were provided by the TOPMed Data Coordinating Center (R01HL-120393; U01HL-120393; contract HHSN268201800001I). We gratefully acknowledge the studies and participants who provided biological samples and data for TOPMed. The Women’s Health Initiative (WHI) program is funded by the National Heart, Lung, and Blood Institute, National Institutes of Health, U.S. Department of Health and Human Services through contracts HHSN268201600018C, HHSN268201600001C, HHSN268201600002C, HHSN268201600003C, and HHSN268201600004C. This manuscript was not prepared in collaboration with investigators of the WHI, and does not necessarily reflect the opinions or views of the WHI investigators, or NHLBI. The Mount Sinai BioMe Biobank is supported by The Andrea and Charles Bronfman Philanthropies.

The All of Us Research Program is supported by the National Institutes of Health, Office of the Director: Regional Medical Centers: 1 OT2 OD026549; 1 OT2 OD026554; 1 OT2 OD026557; 1 OT2 OD026556; 1 OT2 OD026550; 1 OT2 OD 026552; 1 OT2 OD026553; 1 OT2 OD026548; 1 OT2 OD026551; 1 OT2 OD026555; IAA #: AOD 16037; Federally Qualified Health Centers: HHSN 263201600085U; Data and Research Center: 5 U2C OD023196; Biobank: 1 U24 OD023121; The Participant Center: U24 OD023176; Participant Technology Systems Center: 1 U24 OD023163; Communications and Engagement: 3 OT2 OD023205; 3 OT2 OD023206; and Community Partners: 1 OT2 OD025277; 3 OT2 OD025315; 1 OT2 OD025337; 1 OT2 OD025276. In addition, the All of Us Research Program would not be possible without the partnership of its participants.

The Genotype-Tissue Expression (GTEx) Project was supported by the Common Fund of the Office of the Director of the National Institutes of Health (commonfund.nih.gov/GTEx). Additional funds were provided by the NCI, NHGRI, NHLBI, NIDA, NIMH, and NINDS. Donors were enrolled at Biospecimen Source Sites funded by NCI\Leidos Biomedical Research, Inc. subcontracts to the National Disease Research Interchange (10XS170), Roswell Park Cancer Institute (10XS171), and Science Care, Inc. (X10S172). The Laboratory, Data Analysis, and Coordinating Center (LDACC) was funded through a contract (HHSN268201000029C) to The Broad Institute, Inc. Biorepository operations were funded through a Leidos Biomedical Research, Inc. subcontract to Van Andel Research Institute (10ST1035). Additional data repository and project management were provided by Leidos Biomedical Research, Inc.(HHSN261200800001E). The Brain Bank was supported by supplements to University of Miami grant DA006227. Statistical Methods development grants were made to the University of Geneva (MH090941 & MH101814), the University of Chicago (MH090951, MH090937, MH101825, & MH101820), the University of North Carolina - Chapel Hill (MH090936), North Carolina State University (MH101819), Harvard University (MH090948), Stanford University (MH101782), Washington University (MH101810), and to the University of Pennsylvania (MH101822). The datasets used for the analyses described in this manuscript were obtained from dbGaP at http://www.ncbi.nlm.nih.gov/gap through dbGaP accession number phs000424.v7.p2.

## Data and code availability

1000 Genomes Data, https://www.internationalgenome.org/data-portal/data-collection/30x-grch38 Database of Genotypes and Phenotypes (dbGaP), NHLBI TOPMed: Women’s Health Initiative (WHI) https://www.ncbi.nlm.nih.gov/projects/gap/cgi-bin/study.cgi?study_id=phs001237.v2.p1 Database of Genotypes and Phenotypes (dbGaP), NHLBI TOPMed - NHGRI CCDG: The BioMe Biobank at Mount Sinai, https://www.ncbi.nlm.nih.gov/projects/gap/cgi-bin/study.cgi?study_id=phs001644.v2.p2

UKB Data Showcase, https://biobank.ctsu.ox.ac.uk/crystal/search.cgi

All of Us Researcher Workbench, https://www.researchallofus.org/data-tools/workbench/

GTEx DNA methylation data, https://www.ncbi.nlm.nih.gov/geo/query/acc.cgi?acc=GSE213478

GTEx normalized expression level data, https://storage.googleapis.com/gtex_analysis_v8/rna_seq_data/GTEx_Analysis_2017-06-05_v8_RNASeQCv1.1.9_gene_tpm.gct.gz

All genotype data generated in the UKB and All of Us cohorts have been returned and will be available through future data releases.

Code utilized for this study is available as follows:

https://github.com/Illumina/ExpansionHunter

https://github.com/bharatij/Global-Ancestry-Assignment

https://github.com/PacificBiosciences/trgt

## Author contributions

C.A.M., B.J., P.G., M.S. and W.L. designed bioinformatics pipelines and performed data analyses. A.M.T. and A.J.S. supervised the project. A.J.S. conceived the study, performed data analyses and drafted the manuscript. All authors reviewed and approved the final draft.

## Notes

### Competing Interest Statement

The authors have declared no competing interest.

### Author Declarations

Collection of the UKB data was approved by the Research Ethics Committee of the UKB obtained under application 32568. All study participants provided informed consent, and the protocols for UKB are overseen by The UKB Ethics Advisory Committee, see https://www.ukbiobank.ac.uk/ethics/.

