## Supplementary Figures for "A phenome-wide association study of tandem repeat variation in 168,554 individuals from the UK Biobank"

### List of Supplementary Tables

**Supplementary Table 1.** List of all TRs genotyped in the UK Biobank.
**Supplementary Table 2.** List of all traits used for PheWAS in the UK Biobank.
**Supplementary Table 3.** Results from PheWAS on autosomes in the UK Biobank.
**Supplementary Table 4.** Results from PheWAS on chrX in the UK Biobank.
**Supplementary Table 5.** Results from causal variant analysis.
**Supplementary Table 6.** Association results at causal loci after excluding individuals carrying TR alleles in the top 5% of the allelic distribution.
**Supplementary Table 7.** Results from replication analysis in the All of Us cohort.
**Supplementary Table 8.** Results of expression QTL analysis in the GTEx cohort.
**Supplementary Table 9.** Results of methylation QTL analysis in the GTEx cohort.

### Supplementary Figures

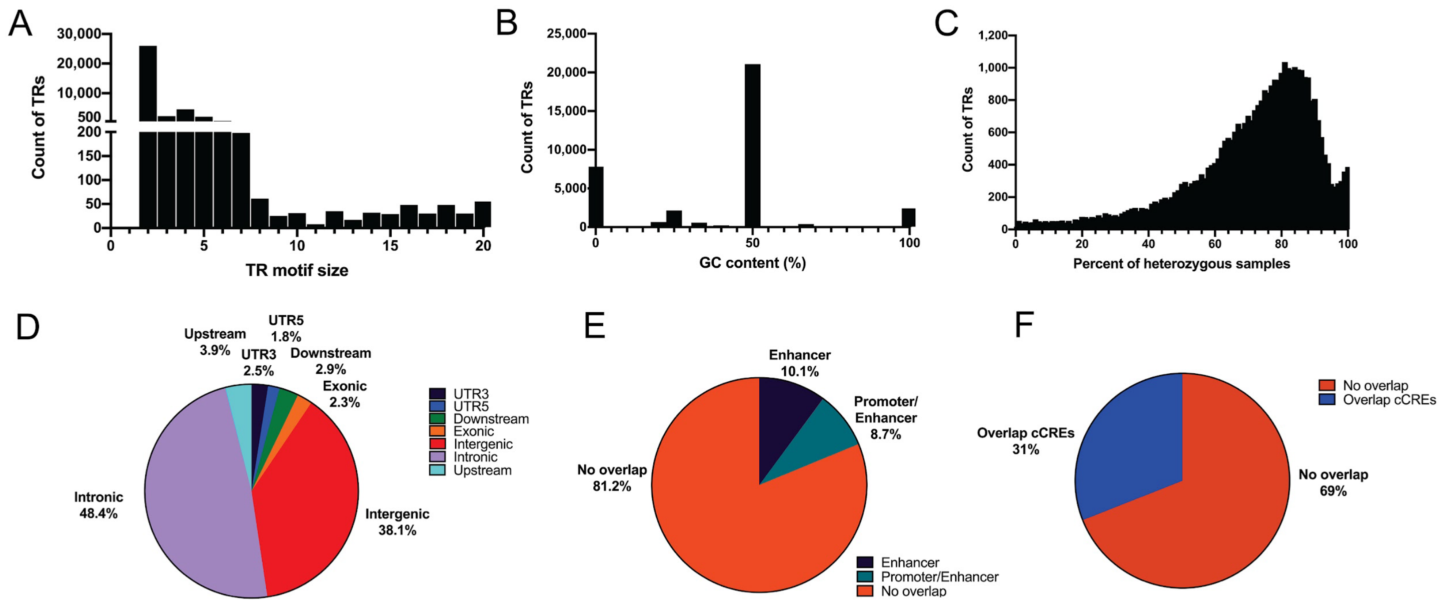


**Supplementary Figure 1. Descriptive statistics of the 36,085 TRs used for PheWAS in the UK Biobank cohort. (A)** Distribution of TR motif sizes. **(B)** Distribution of GC-content of TR motifs. **(C)** Distribution of heterozygosity rates in the UK Biobank cohort. **(D)** Overlap with genic annotations. **(E)** Overlap with GeneHancer regulatory annotations. **(F)** Overlap with ENCODE candidate *cis* regulatory elements. In D and E, for clarity, we do not display categories that are ≤0.5%.

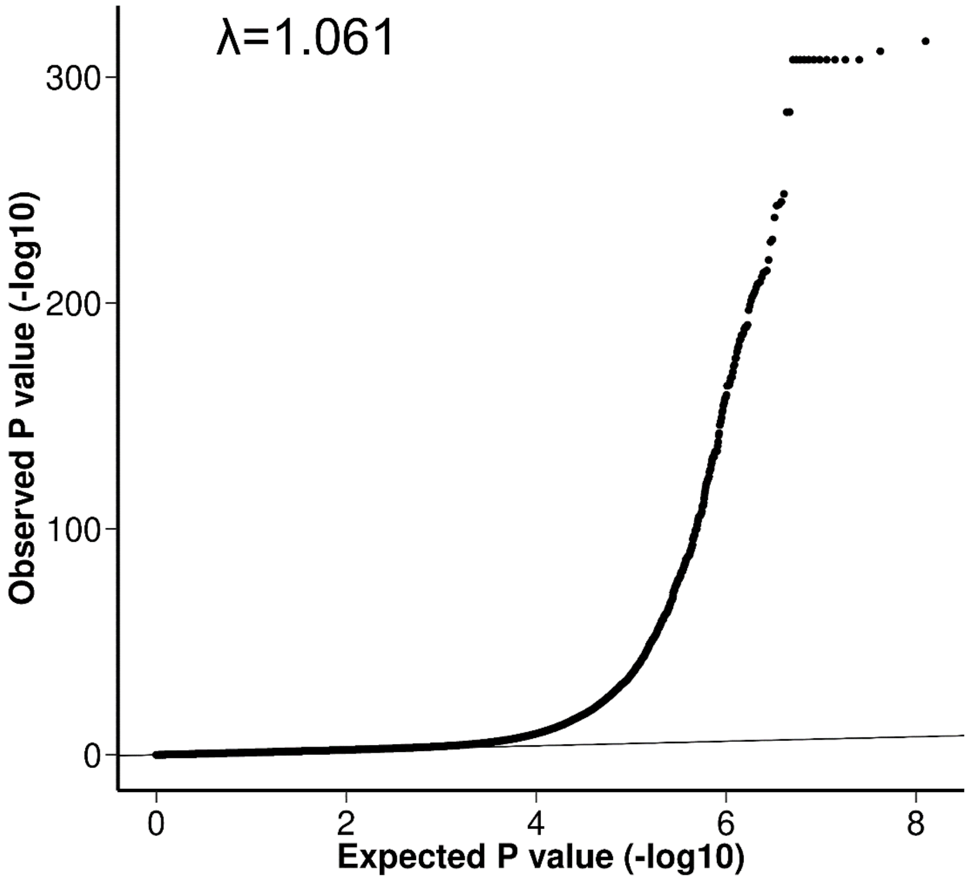


**Supplementary Figure 2. QQ plot based on associations with autosomal TRs.** We observed a strong excess of significant signals compared to the null expectation. Genomic inflation was well controlled, with an inflation value (λ) of 1.061.

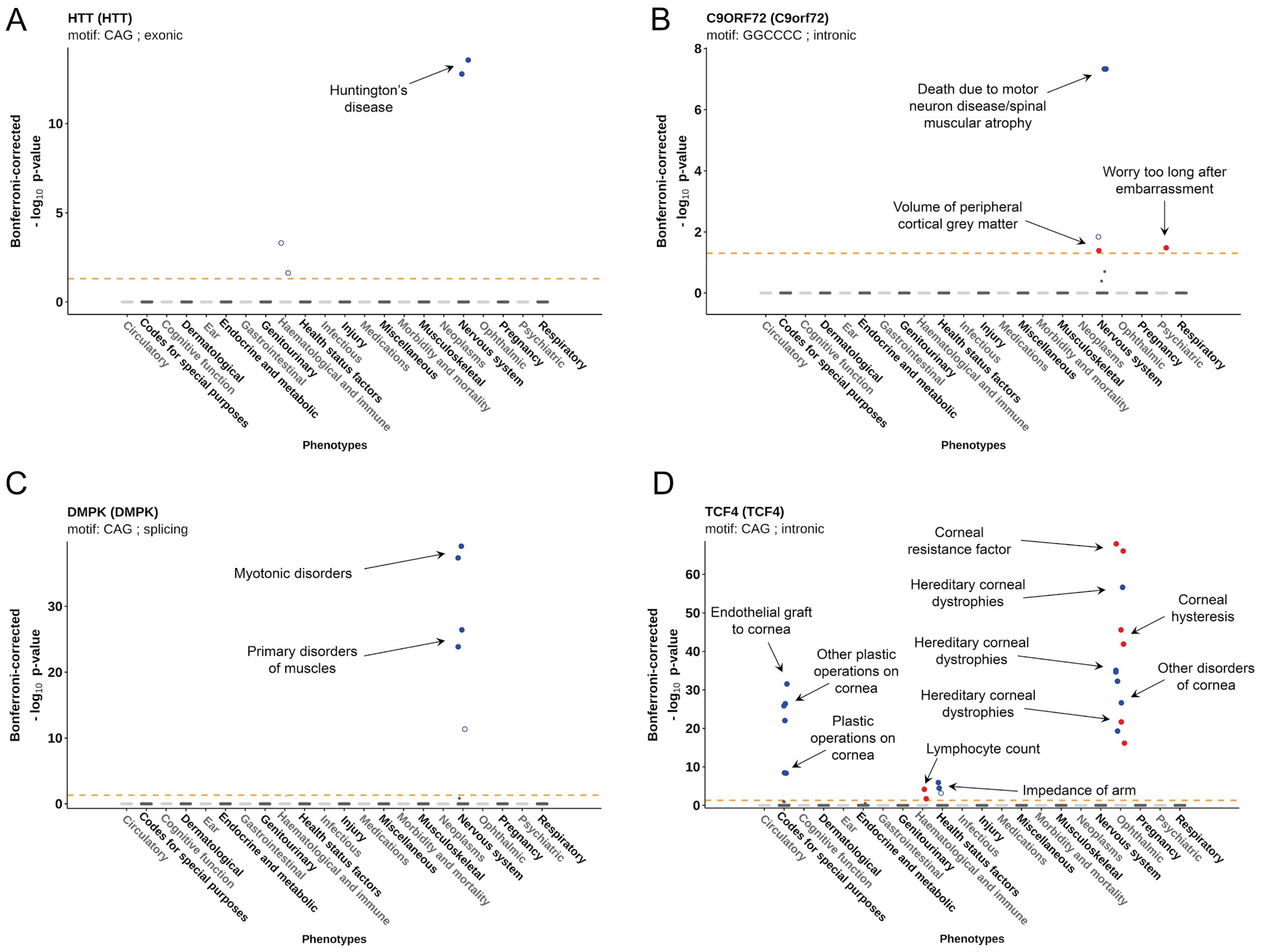


**Supplementary Figure 3. Causal associations with four known repeat expansion disorder loci recapitulate the known effects of these diseases.** In each case, the associated traits we identified by PheWAS were consistent with the known phenotypic effects of extreme expansions of these TRs. In each plot, traits are grouped into physiological categories (x-axis), and we plot the -$\log_{10}$ Bonferroni-corrected p-value (y-axis). Solid filled points indicate where the TR was scored as a high-confident causal association, with blue for positive direction of effect and red for negative direction of effect. Unfilled points indicate associations that were not considered causal. The horizontal dashed line indicates the Bonferroni-corrected threshold of $p<0.05$.

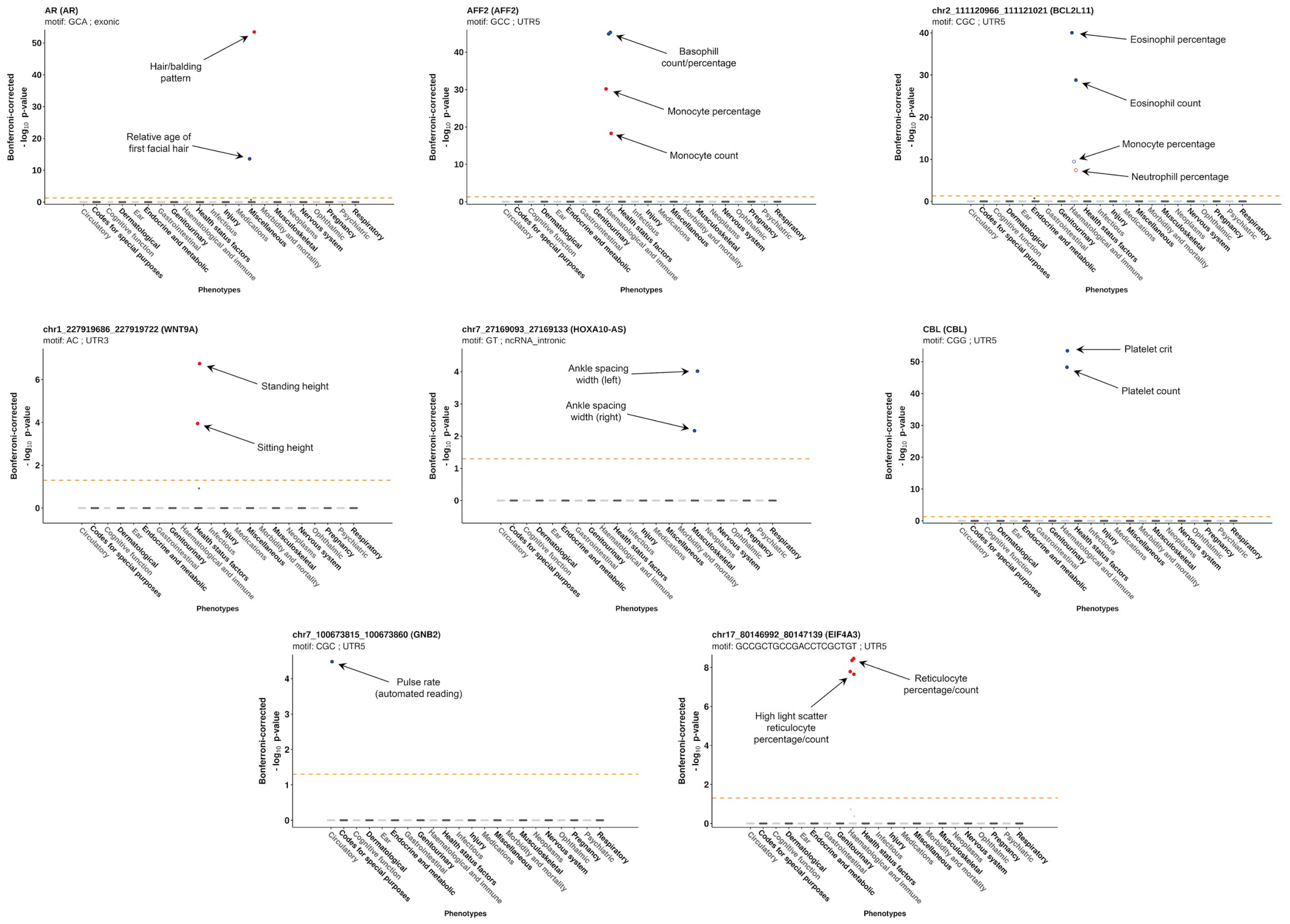


**Supplementary Figure 4. Examples of causal associations identified with common polymorphic variation in TRs.** In each plot, traits are grouped into physiological categories (x-axis), and we plot the -$\log_{10}$ Bonferroni-corrected p-value (y-axis). Solid filled points indicate where the TR was scored as a high-confident causal association, with blue for positive direction of effect and red for negative direction of effect. Unfilled points indicate associations that were not considered causal. The horizontal dashed line indicates the Bonferroni-corrected threshold of $p<0.05$.

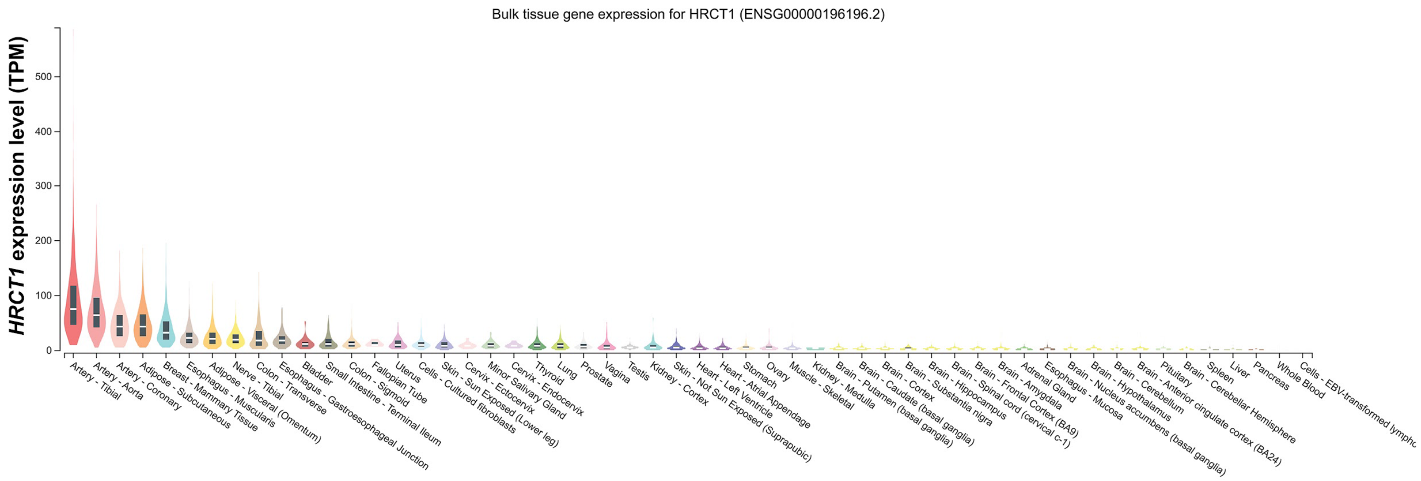


**Supplementary Figure 5.** *HRCT1*, a gene of unknown function, shows highest expression in arterial tissue out of 48 tissues analyzed by the GTEx cohort.

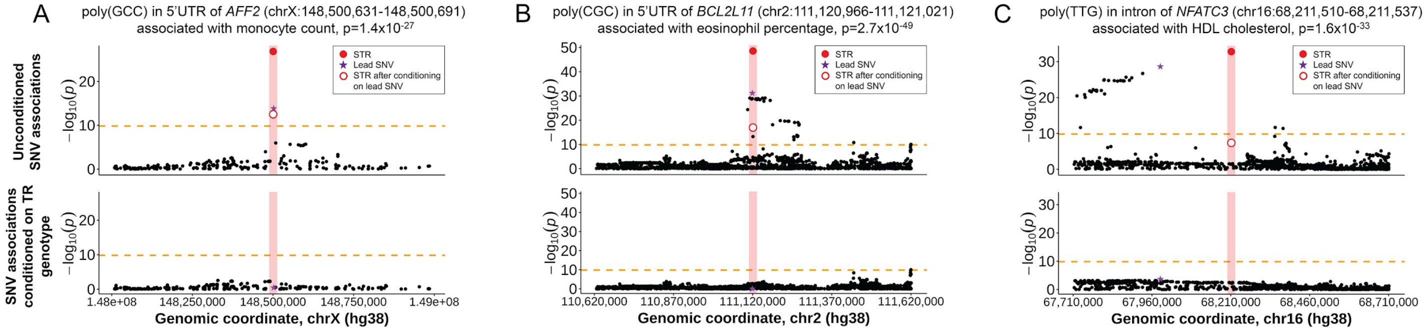


**Supplementary Figure 6. Results of conditional analysis at high-confidence causal loci.** For each locus, the upper plot shows the original association signals for the TR (red filled circle) and SNVs within ±500kb (black points), in addition to the association signal for the TR after conditioning (unfilled red circle) based on the genotype of the lead SNV at the locus (purple star). The lower plots show the association signals for the same set of SNVs after conditioning each one based on the average genotype of the causal TR. In each plot, the shaded vertical red bar shows the position of the causal TR, while the horizontal dashed line indicates the Bonferroni significance threshold of $p<1.45\times10^{-10}$.
